# ERAP1, ERAP2, and two copies of HLA-Aw19 alleles increase the risk for Birdshot Chorioretinopathy in HLA-A29 carriers

**DOI:** 10.1101/2021.07.02.21259921

**Authors:** Sahar Gelfman, Dominique Monnet, Ann J. Ligocki, Thierry Tabary, Arden Moscati, Xiaodong Bai, Jan Freudenberg, Blerta Cooper, Jack A. Kosmicki, Sarah Wolf, Regeneron Genetics Center, Manuel A. R. Ferreira, John Overton, Jonathan Weyne, Eli A. Stahl, Aris Baras, Carmelo Romano, Jacques H. M. Cohen, Giovanni Coppola, Antoine Brézin

**Affiliations:** Regeneron Genetics Center, 777 Old Saw Mill River Rd., Tarrytown, NY 10591, USA; Université de Paris, Hôpital Cochin, service d’ophtalmologie, Paris, France; Regeneron Pharmaceuticals, 777 Old Saw Mill River Rd., Tarrytown, NY 10591, USA; University of Reims Champagne Ardennes, LRN EA4682, 51092 Reims, France

## Abstract

**Purpose:** Birdshot Chorioretinopathy (BSCR) is strongly associated with HLA-A29. This study was designed to elucidate the genetic modifiers of BSCR in HLA-A29 carriers.

**Methods:** We sequenced the largest BSCR cohort to date, including 286 cases and 108 HLA-A29 positive controls to perform genome wide common and rare variant associations. We further typed the HLA alleles of cases and 45,386 HLA-A29 controls of European ancestry to identify HLA alleles that associate with BSCR risk.

**Results:** Carrying a second allele that belongs to the HLA-Aw19 broad antigen family (including HLA-A29, A30, A31, and A33) increases the risk for BSCR (OR=4.44, p=2.2e-03). This result was validated by comparing allele frequencies to large HLA-A29-controlled cohorts (n=45,386, OR>2.5, p<1.3e-06). We also confirm that ERAP1 and ERAP2 haplotypes modulate the risk for disease within our HLA-A29 controlled cohort. A meta-analysis with an independent dataset confirmed that ERAP1 and ERAP2 haplotypes modulate the risk for disease at a genome-wide significant level: ERAP1-rs27432 (OR 2.46; 95% CI 1.85-3.26; p=4.07e-10), an eQTL decreasing ERAP1 expression, and ERAP2-rs10044354 (OR 1.95; 95% CI 1.55-2.44; p=6.2e-09), an eQTL increasing ERAP2 expression. Furthermore, ERAP2-rs2248374 that disrupts ERAP2 expression is protective (OR 0.56; 95% CI [0.45-0.70]; p=2.39e-07). BSCR risk is additively increased when combining ERAP1/ERAP2 risk genotypes with two copies of HLA-Aw19 alleles (OR 13.53; 95% CI 3.79-54.77, p=1.17e-05).

**Conclusions:** The genetic factors increasing BSCR risk demonstrate a pattern of increased processing, as well as increased presentation of ERAP2 specific peptides. This suggests a mechanism in which exceeding a peptide presentation threshold activates the immune response in choroids of A29 carriers.

## Introduction

BSCR is a rare bilateral posterior uveitis affecting individuals of European descent with a mean age of 53 years at the time of diagnosis, characterized by a mild vitritis and ovoid cream-colored lesions at the level of the choroid ^1-4^. The retinal inflammation can include vasculitis, papillitis and macular edema. Greater than 95% of BSCR patients carry the HLA-A29 allele, corresponding to an OR of 157.5, the strongest known HLA class I association with any disease ^5-9^. Indeed, some investigators believe that the diagnosis of the disease can only be made in HLA-A29 carriers^10^. Histologic analyses of eyes with BCR have revealed nongranulomatous nodular infiltrations of the choroid and lymphocytic infiltrates ^11,12^. BSCR can result in a severe gradual loss of vision and is usually treated by immunosuppressants or immunomodulating biological therapies.

Despite the association with an HLA class-I protein, expressed on most nucleated cells, the pathology of BSCR is restricted to the posterior ocular tissues. Additionally, the number of estimated BSCR cases in the US (5,000-10,000) is far lower than the HLA-A29 prevalence (7%) in European population ^5,9^, suggesting that HLA-A29 is necessary but not sufficient to cause disease, and that other factor(s) contribute to the development of BSCR. Previous work identified risk haplotypes and polymorphisms in both *ERAP1* (rs2287987, Hap10) and *ERAP2* (rs10044354, HapA) that are more prevalent in BSCR patients relative to healthy controls ^8,13^. ERAP1 and ERAP2 are endoplasmic reticulum aminopeptidases with complementary substrate preferences that trim peptides to be loaded onto, and presented by, HLA class I proteins ^14^. Furthermore, expression of ERAP1 and ERAP2 is coordinated, and when ERAP1 expression is decreased, ERAP2 expression is increased ^15^. Alterations in the expression or enzymatic activity levels of ERAP1 and ERAP2 have been shown to alter the peptidome available for presentation by HLA class I proteins^16,17^. The *ERAP1* risk associated Hap10 polymorphism is associated with lower expression, as well as reduced peptide trimming activity, of the ERAP1 protein^14^. The *ERAP2* risk associated HapA polymorphism produces a full-length functional ERAP2 protein whereas the protective HapB polymorphism produces no detectable full-length ERAP2 protein ^14,18^. The combination of a hypoactive ERAP1 and an available ERAP2 is therefore hypothesized to lead to a unique peptidome pool available for presentation by HLA class I proteins. Variation in the canonical peptide motifs in the peptide-binding groove of the HLA class I molecules^16,19^ further contributes to shaping the peptidome present on the cell surface for immune cell interrogation, recognition, and subsequent immune activation in BSCR.

Here we study the largest BSCR cohort, consisting of 286 cases and 108 HLA-A29 controls of European ancestry obtained by the University of Paris (UParis cohort). We investigated the effect of other HLA-A alleles in a HLA-A29-controlled cohort and identified a novel independent risk associated with most members of the Aw19 serotype family (HLA-A29 homozygous, A30, A31, or A33), while one member, HLA-A32, is depleted in BSCR patients compared to A29 positive controls. Our analyses also confirmed the BSCR risk associations with both *ERAP1* Hap10 and *ERAP2* HapA haplotypes. Our results shed light on the underlying mechanism that support the activation of the immune response in A29-positive individuals and resulting in BSCR.

## Methods

### Study subjects and samples

The genomic DNA samples from 286 patients with BSCR and 108 unrelated healthy local French volunteers that exhibited HLA tissue typing common in the French population were included in this study. The patients were recruited at Hôpital Cochin, Paris, France. All patients met the criteria for diagnosis of BSCR as defined both by an international consensus conference held in 2002 and by the Standardization of Uveitis Nomenclature (SUN) Working Group ^6,20^. In brief, all patients had a posterior bilateral uveitis with multifocal cream-colored or yellow-orange, oval or round choroidal lesions (“birdshot spots”). Although the presence of the HLA-A*29 allele was not a requirement for the diagnosis of BSCR according to the international criteria, all patients included in the current study carried the HLA-A*29 allele. The control DNA samples were collected from volunteer donors recruited by the hematopoietic stem cell donor center of Rheims for France Greffe de Moelle Registry, and local control healthy individuals of the Registry. The DNA samples were isolated from peripheral blood samples using a standard salting out method or QIAamp Blood Kit (Qiagen, Chatsworth, CA, USA). Quality and quantity of DNA was determined by UV spectrophotometry and the concentration was adjusted to 100 ng/ml. Signed informed consent documentation was obtained from all participants, and all research adhered to the tenets set forth in the Declaration of Helsinki. All study-related data acquisitions were approved by the Paris Cochin institutional review board.

### Genetic data

We took a comprehensive approach to both sequence the exomes and genotype all samples, to allow for identification of common and rare variants filtered based on high quality calls. DNA from participants was genotyped on the Illumina Global Screening Array (GSA) and imputed to the HRC reference panel. Prior to imputation, we retained variants that had a MAF >= 0.1%, missingness < 1% and HWE p-value > 10^−15^. Imputation using the HRC reference panel yielded 8,385,561 variants with imputation INFO>0.3 and MAF>0.5%.

Exome sequencing was performed to a mean depth of 31x, followed by variant calling and quality control as reported previously^21^, resulting in 238,942 variants. When integrated, this produced an overall dataset with 8,459,907 variants: 65.5% common (MAF > 5%), 34.5% low-frequency (0.5% < MAF < 5%) and 0.01% rare (MAF < 0.5%).

### HLA genotyping

HLA Class I genes (HLA-A, -B, and -C) were amplified in a multiplex PCR reaction with primers encompassing the full genomic loci for each target. The resulting amplicons were enzymatically fragmented to an average size of 250 base pairs and prepared for Illumina sequencing (New England Biolabs, Ipswich, MA). The libraries were sequenced on the Illumina HiSeq 2500 platform on a rapid run flow cell using paired-end 125 base pair reads with dual 10 base pair indexes. Upon completion of sequencing, raw data from each Illumina HiSeq run was gathered in local buffer storage and uploaded to the DNAnexus platform^22^ for automated analysis. The FASTQ-formatted reads were converted from the BCL files and assigned to samples identified by specific barcodes using the bcl2fastq conversion software (Illumina Inc., San Diego, CA). All the reads in sample-specific FASTQ files were subject to HLA typing analysis using an updated version of PHLAT program^23^ with the reference sequences consisting of GRCh38 genomic sequences and HLA type reference sequences in the IPD-IMGT/HLA database v3.30.0^24^.

In addition, HLA allele imputation was performed following SNP2HLA^25^ with the T1DGC HLA allele reference panel^26^. HRC-imputed genotypes in the extended Major Histocompatibility Complex (MHC) region (chr6:25-35Mb) were filtered for high INFO score (>0.9) and certainty (maximum GP>0.8 for all genotyped), in order to increase overlap with the T1DGC reference panel, were re-phased along with chromosome 6 array genotypes using SHAPEIT4^27^, and were imputed using Minimac4^28^. HLA allele imputation quality was assessed by examining INFO score vs MAF, and imputed vs reference panel MAF.

### Genetic association analyses

Association analyses in each study were performed using the genome-wide Firth logistic regression test implemented in SAIGE ^29,30^. In this implementation, Firth’s approach is applied when the p-value from standard logistic regression score test is below 0.05. We included for the GRM for SAIGE directly genotyped variants with a minor allele frequency (MAF) >1%, <10% missingness, Hardy-Weinberg equilibrium test *P*-value>10^−15^ and linkage-disequilibrium (LD) pruning (1000 variant windows, 100 variant sliding windows and *r*^2^<0.1). The association model included as covariates sex and the first 10 ancestry-informative principal components (PCs) derived from the GRM dataset. Haplotype analyses were performed using PLINK 1.0^31^ *--chap* and *--hap-assoc* and --*hap-logistic*, and in *R*. High haplotype imputation and phasing quality was indicated by PLINK *--hap-phase* maximum likelihood haplotype genotypes’ posterior probabilities all equal to one.

### HLA-A allele association analyses

Association of HLA-A alleles was performed as follows: for each sample, we typed both HLA-A alleles as described above. Following HLA allele typing, we removed related samples. For the remaining cohort of 282 cases and 106 controls, we next obtained one HLA-A allele that is not A29 (the “second” allele). Samples carrying two copies of A29, we considered A29 as the second allele. We then subjected the cohort to a Fisher’s exact test, in which we tested the association of each allele that was identified in three or more BSCR cases, with the case-control status. To answer the question of whether the A19 allele group is also associated with the case-control status, we combined, and tested together the samples in two different ways: carrying all Aw19 alleles (A29, A30, A31, A32 and A33). Since A32 is biologically different than the other Aw19 alleles in its peptide binding domain^32^, we also constructed and tested a group that is made of samples carrying all Aw19 alleles excluding A32. The final odds-ratios and p-values are presented in Table 1.

**Table 1.**
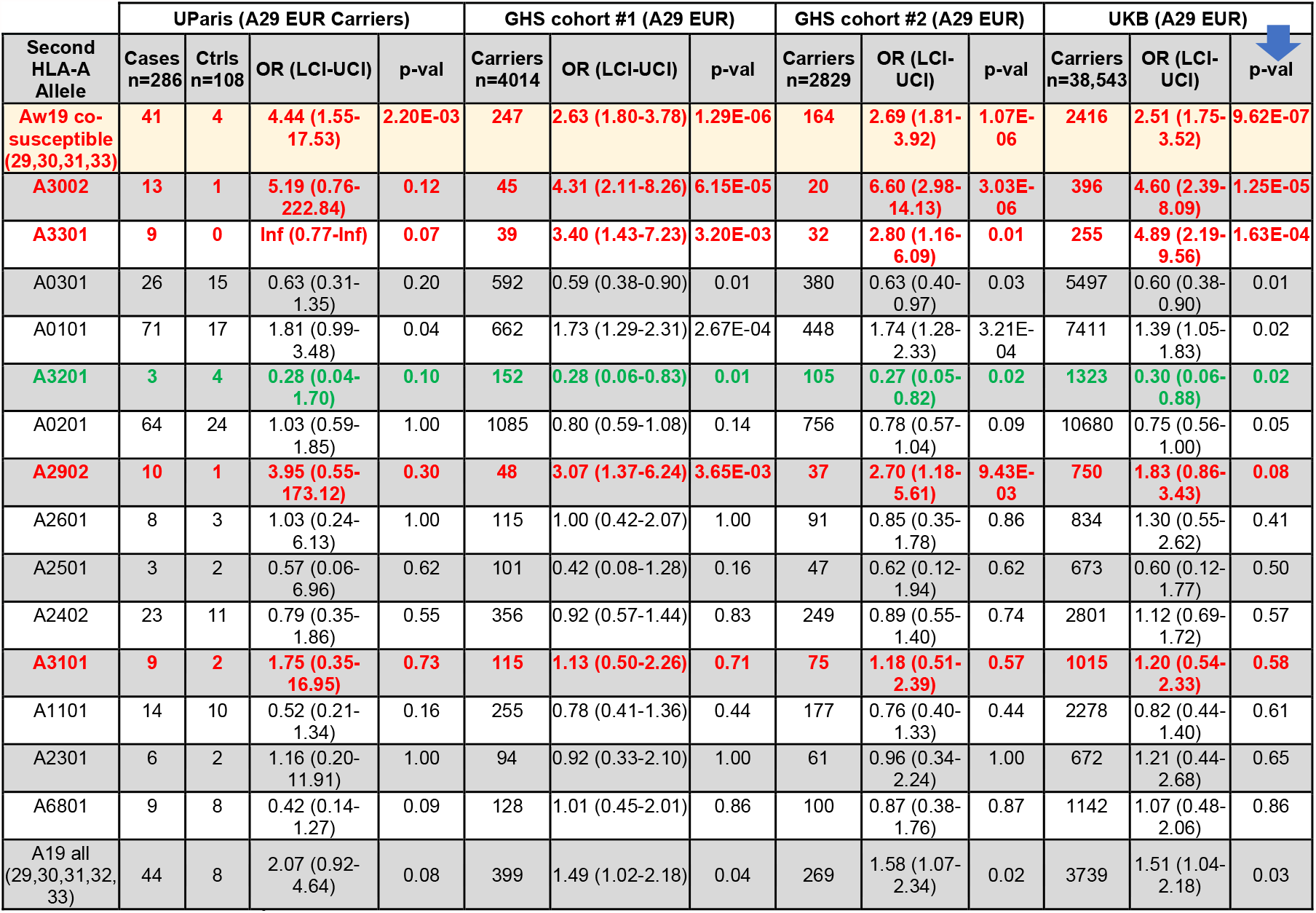
HLA-A 2^nd^ allele frequencies in the French cohort compared to UKB and GHS EUR A29 carriers. Alleles belonging to the Aw19 broad antigen group that increase risk are A29, A30, A31 and A33 (red) and A32 exhibits protection (green). A Fisher’s exact test combining all Aw19 risk alleles presents the strongest enrichment in all comparisons. Only alleles that have three or more case carriers are presented. Table is sorted by p-values when comparing case frequencies against A29 controls in UKB.

## Results

### HLA-Aw19 broad antigen serotype alleles and BSCR risk

Our HLA-A29-controlled cohort allowed us to examine the HLA region while controlling for the strong association of HLA-A29 with BSCR, and therefore to detect possible additional association signals in the HLA region.

First, we asked whether rare variants on the HLA-A29 background were enriched in BSCR cases. We did not identify significant enrichments [of rare single or aggregated variants] either within or outside the MHC region.

Second, we asked whether other HLA-A alleles in addition to the HLA-A29 allele increased BSCR risk. We constructed an assay to type HLA-A alleles in this cohort (see Methods), and tested the second HLA-A allele (other than the known first HLA-A29) for association with BSCR. We found that additional HLA-A alleles were associated with BSCR, and those with the largest effects belonged to the same HLA-Aw19 broad antigen serotype group: HLA-A29:02, A30:02, A31:01 and A33:01 (Table 1). As a group, HLA-Aw19 alleles were significantly enriched in the second allele of BSCR patients (OR=4.44, p=2.2e-03, figure 1, blue bars). This result suggests, for example, that individuals carrying two copies of HLA-A29 would be at a greater risk of developing BSCR compared to those carrying one copy. It also suggests that other Aw19 allele may play a role in BSCR co-susceptibility or pathogenesis in concert with A29. The sole exception within the HLA-Aw19 serotype group is HLA-A32, which has been reported not to share the defining Aw19 binding domain^32^; HLA-A32 appears to be depleted in BSCR cases and thus protective against BSCR (OR=0.28, p=0.1).

**Figure 1.**
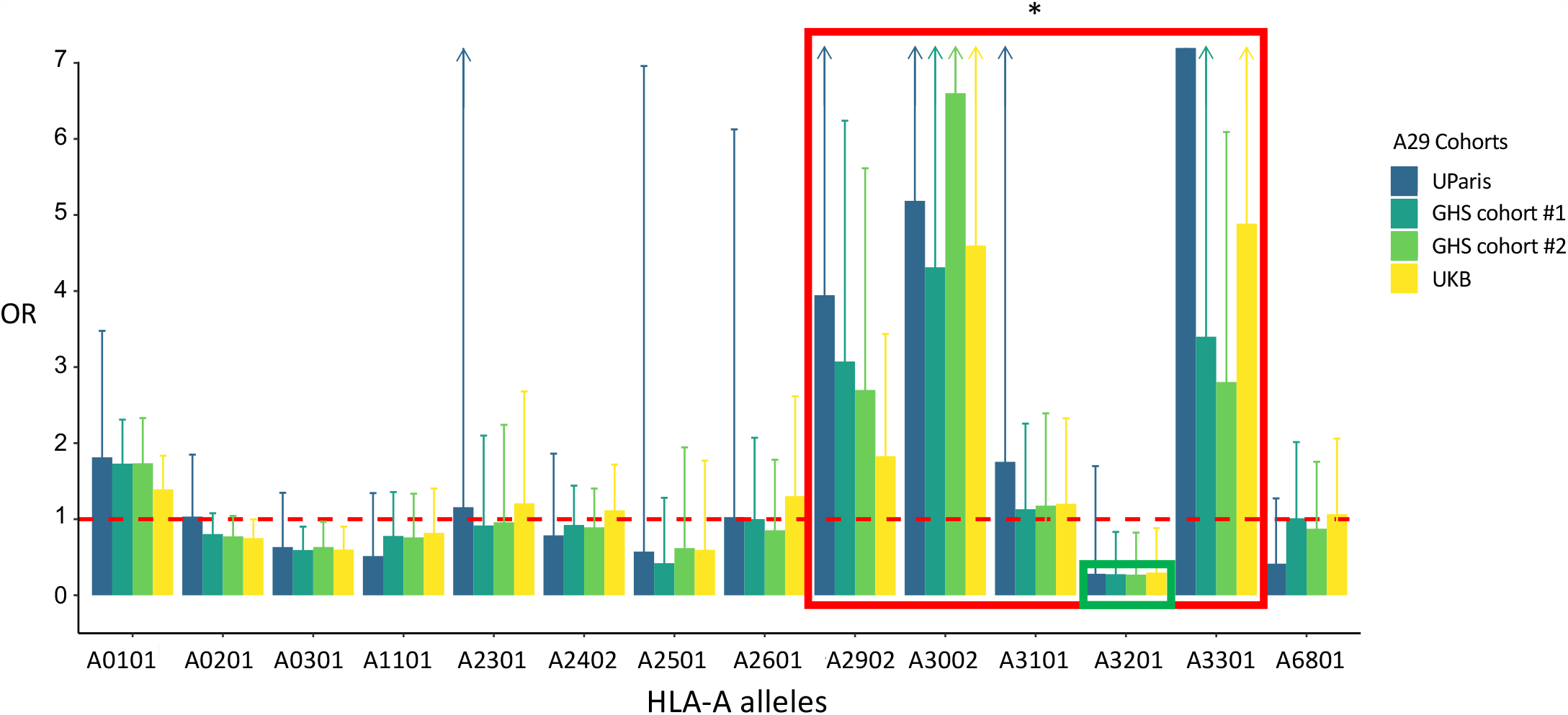
Aw19 enrichment in Birdshot cases. Odds-ratio for BSCR, comparing frequencies of 14 HLA-A alleles that are present in three or more cases (>1%, x-axis) in 286 UParis cases compared with 108 UParis controls (Blue), GHS control cohort #1 (n= 4,014, dark green), GHS control cohort #2 (n= 2,829, bright green) and UKB controls (n= 38,543, yellow). Aw19 alleles show the highest ORs (red box) that replicates with large A29 control cohorts, with the exception of A32 that is depleted in cases (green box). * p<0.01

**Figure 2.**
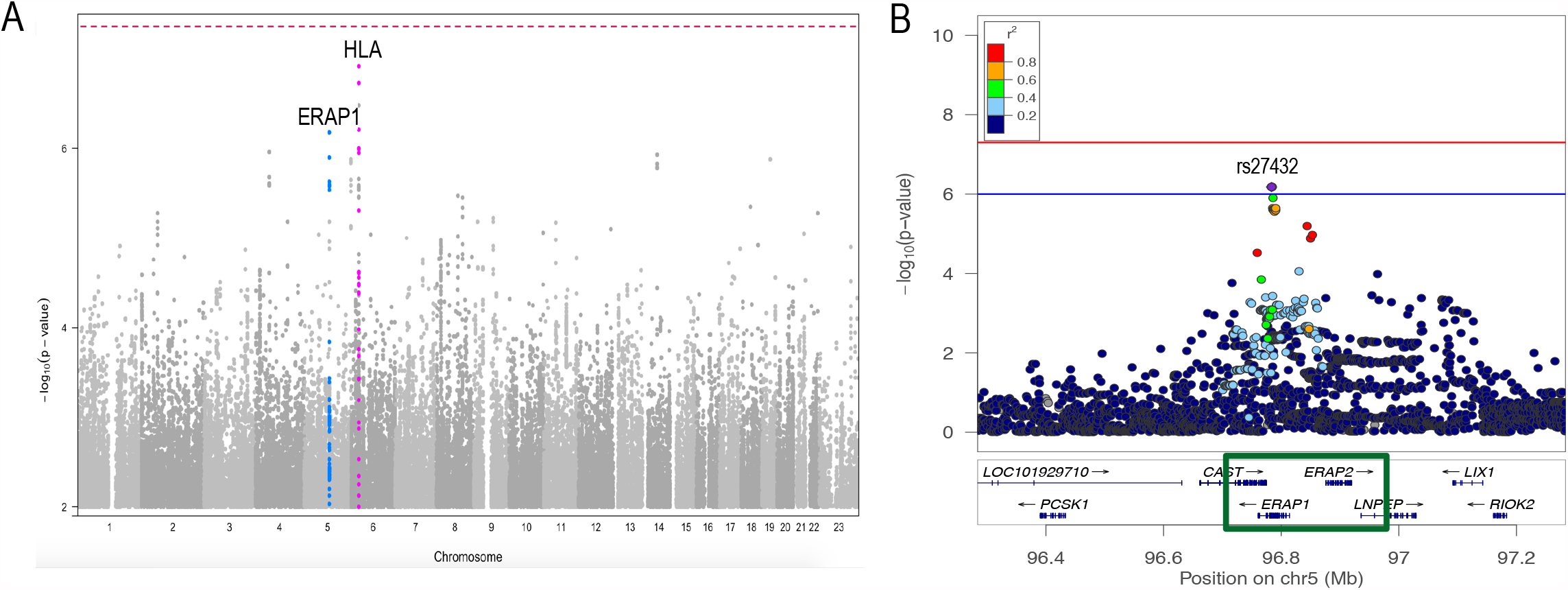
Manhattan and ERAP1 locus zoom plot of the A29-stratified French cohort. **A)** Birdshot association analysis of 286 A29 cases and 108 A29 controls showing borderline associations (p<1e-6) at several loci including HLA on chromosome six (pink) and ERAP1 on chromosome five (blue). **B)** Locus zoom plot exhibiting the *ERAP1-ERAP2* locus and the top *ERAP1*-rs27432 risk variant.

The above results presented two issues due to the small numbers of controls in UParis (n=108): 1) The frequency of alleles might not represent the frequency of HLA-A alleles in general EUR population. 2) While the high ORs replicate in several HLA-Aw19 alleles, the numbers are not sufficient to support significant associations. To tackle these concerns, we examined the frequency of HLA-A alleles in three other large European (EUR) ancestry control populations, two cohorts from the Geisinger Health System (GHS cohort #1, n=77,198 and GHS cohort #2, n=59,072) and the UK Biobank (UKB, n=463,315). In all three datasets, we selected EUR samples carrying at least one HLA-A29 allele, matching our BSCR cohort: 4,014 A29 carriers from GHS cohort #1 (5.2% of all EUR subjects), 2,829 A29 carriers from GHS cohort #2 (4.8% of all EUR), and 38,543 A29 carriers from UKB (8.3% of all EUR). We compared the frequencies of the second HLA-A alleles in these cohorts to those observed in our BSCR cohort (Figure 1, Table 1). The results support the enrichment of four of the five HLA-Aw19 alleles in BSCR cases, with highest increased risk for HLA-A30:02 (GHS cohort #1 OR=4.31, GHS cohort #2 OR=6.6, UKB OR= 4.6) and HLA-A33 (GHS cohort #1 OR=3.4, GHS cohort #2 OR=2.8, UKB OR= 4.9). When combining samples carrying the four co-susceptibility alleles A29, A30, A31 and A33, we found a highly significant enrichment in BSCR cases when compared with the larger control cohorts (GHS cohort #1 p-val= 1.29E-06, GHS cohort #2 p-val= 1.07E-06, UKB p-val= 9.62E-07, Table 1 top row). This analysis excludes A32 because of its biological difference in the sequence of the peptide binding domain as previously reported^32^ (see Discussion). That said, we performed additional analyses with all Aw19 alleles including A32, and we find that the enrichment in cases is reduced when it is included (Table 1 bottom row). We also tested whether these associations are affected by measurable confounders, conducting logistic regression tests to evaluate the effects of the second HLA-A allele in HLA-A29 carriers, in UParis BSCR cases compared with each control cohort, with covariates included for sex and principal components, calculated based on genetic array data for each analytic set (supplementary table 1). We found that these results were consistent with increased risk for the HLA-Aw19 co-susceptibility alleles, A29, A30, A31, and A33.

### HLA-A32 exhibits protection from BSCR in an HLA-A29 positive cohort

HLA-A32 is underrepresented in BSCR cases (3/286, ∼1%) versus A29 carrier controls (4/108, 3.7%), corresponding to a nominally significant protection from risk (OR=0.28, p=0.1; Table 1). When compared with the larger control cohorts, the trend protection is maintained with both UKB controls (3.4%, OR=0.3, p=0.02) and GHS controls (cohort #1: 3.8%, OR=0.27, p=0.01; cohort #2: 3.7%, OR=0.27, p=0.02). While nominally significant, this result does not pass the threshold of multiple test correction (p=3.57e-03) and will need to be further validated with additional case cohorts.

### *ERAP1* and *ERAP2* are Independently Associated with BSCR

We tested all variants and gene burdens for association with case-control status, while controlling for sex and ten principal components, using a generalized linear mixed model (SAIGE^8^, see Methods). Due to the fact that both cases and controls were A29 allele carriers, the expected strong HLA-A signal was at least partially controlled, as evidenced by the strongest HLA p-value = 8.98E-07, compared with p=6.6e-74 with 125 cases in the previous BSCR report ^8^. Overall, no locus passed the genome wide significance threshold (p< 5e-8). Other than the remnant signal at HLA-A, only the *ERAP1/ERAP2-LNPEP* locus on chromosome 5 showed an association with disease at p<1e-6 (Figure 1).

The top association within the *ERAP1/ERAP2-LNPEP* locus is the *ERAP1* intronic variant rs27432 (OR [95% CI]= 2.58 [1.78 – 3.76], p= 6.6e-7), a strong eQTL associated with decreased *ERAP1* expression^13,15^, which also tags the risk-increasing common *ERAP1* haplotype^8^. We performed a comparable analysis to assess *ERAP1* haplotype associations with BSCR status in our data. The results are consistent with three levels of risk differentiated by nonsynonymous *ERAP1* variant haplotypes corresponding to Kuiper et al.^15^ Haps 1+2 (OR=0.41, Cases AF=0.17, controls AF=0.35, p=6.7e-06), Hap10 (OR=1.78, Cases AF=0.28, controls AF=0.17, p=8.0e-03), and haplotypes 3-8 (OR=1.32, Cases AF=0.55, controls AF=0.48, p=0.11) (supplementary table 2).

The previously reported top association for BSCR at this locus tags a common variant near ERAP2/LNPEP, rs10044354^8^. This reported risk allele is in a strong linkage disequilibrium (D’=0.99, R^2^=0.76) with a strong eQTL increasing *ERAP2* expression^8^. Our results show a nominal association of rs10044354 with increased risk for Birdshot (OR [95% CI] = 1.55 [1.13 - 2.11], p= 5.8e-3). Furthermore, we did not find significant evidence for an interaction of rs10044354 with rs27432-rs2287987 haplotypes (conditional haplotype test p=0.46).

We next performed a meta-analysis of our results with the published results from Kuiper et al.^8^ which yielded genome-wide significant associations for both *ERAP1* (rs27432, OR [95% CI]=2.46 [1.85-3.26], p=4.07e-10) and *ERAP2* (rs10044354, OR[95% CI]=1.95 [1.55-2.44], p=6.2e-09) loci with BSCR (Table 1). Both previous and current studies showed consistent directionality for both variants, which, separated by over 201,222 bp, show low linkage disequilibrium (LD) in our cohort (R2=0.18, D’=0.79).

The expression of *ERAP2* has been previously reported to be disrupted by a common splice region variant (rs2248374, AF=0.53) that causes mis-splicing of intron 10 and eventual transcript degradation via nonsense-mediated decay ^18,33^, and which is in high LD with rs10044354 (R2=0.8, D’=1). Thus, ∼25% of the population of most ancestries (including European, AF=0.53; African, AF=0.57 and South Asian, AF=0.58) is estimated to be lacking an active ERAP2 protein. We examined both datasets for rs2248374 associations and found that it is protective for BSCR with nominal significance in both datasets (Table 2). In summary, higher expression of ERAP2 protein increases risk for BSCR and a lower expression is protective.

**Table 2.** Top SNPs in *ERAP1* and *ERAP2* regions. Variants in ERAP1 and ERAP2 are genome-wide significant when analyzed together with previous results (125 cases and 670 controls (Kuiper 2014). Rs10044354 is the top association in the *ERAP1-ERAP2* locus in the previous GWAS of Dutch and Spanish cohorts, while rs27432 is the top association in the region in the current French cohort. The LD between the two loci is also presented. *The reference A-allele is the minor allele, risk is the G-allele

| Gene | Variant | Variant type | Study | OR (LCI-UCI) | p-value | Hom OR | Case MAF | Control MAF | Meta OR (LCI-UCI) | Meta p-value |
| --- | --- | --- | --- | --- | --- | --- | --- | --- | --- | --- |
| ERAP2-LNPEP | rs10044354<br>5:96984791:C:T | Intronic | Kuiper et al. | **2.3**<br>(1.69-3.61) | 1.21E-06 |  | 0.63 | 0.42 | **1.95**<br>(1.55-2.44) | **6.20E-09** |
|  |  |  | UParis | **1.55**<br>(1.13-2.11) | 5.80E-03 | **2.6**<br>(1.3-5.15) | 0.52 | 0.41 |  |  |
| LD between rs10044354 (ERAP2-LNPEP) and rs27432 (ERAP1):<br>D' = 0.79 R2 = 0.18 |  |  |  |  |  |  |  |  |  |  |
| ERAP1 | rs27432<br>5:96783569:A:G* | Intronic | Kuiper et al. | **2.26**<br>(2.05-2.47) | 7.20E-05 |  | 0.85 | 0.71 | **2.46**<br>(1.85-3.26) | **4.07E-10** |
|  |  |  | UParis | **2.58**<br>(1.78-3.76) | 6.60E-07 | **4.77**<br>(1.98-11.51) | 0.83 | 0.65 |  |  |

**Table 3.** ERAP2 splice region variant is protective for BSCR. The common *ERAP2* splice region variant rs2248374 that disrupts *ERAP2* expression is protective in the current BSCR cohort and the previous Spanish and Dutch cohorts.

| Gene | Variant | Variant type | Study | OR (LCI-UCI) | p-value | Hom OR | Case MAF | Control MAF | Meta OR (LCI-UCI) | Meta p-value |
| --- | --- | --- | --- | --- | --- | --- | --- | --- | --- | --- |
| ERAP2 | rs2248374<br>5:96900192:A:G | Splice region | Kuiper et al. | **0.44**<br>(0.31-0.63) | 6.60E-06 |  | 0.33 | 0.53 | **0.56**<br>(0.45-0.70) | **2.39E-07** |
|  |  |  | UParis | **0.68**<br>(0.5-0.92) | 1.40E-02 | **0.45**<br>(0.23-0.87) | 0.43 | 0.53 |  |  |

### Cumulative effect of HLA-Aw19 alleles and *ERAP1/ERAP2* haplotypes on BSCR risk

We next examined potential interactions between the *ERAP1* and *ERAP2* association signals, and between HLA-Aw19 and *ERAP1/ERAP2* signals, by calculating the cumulative effects of HLA-Aw19, ERAP1 and ERAP2 genotypes on BSCR risk using the 286 cases and the 4,014 A29 carriers from the GHS cohort #1. First, we performed an analysis of *ERAP2*-rs10044354 risk haplotype, the top non-MHC signal in Kuiper et al.^8^, stratified by single (A29/-) versus double (A29/AW19) Aw19 background, which yielded a trend of increased risk with additional *ERAP2*-rs10044354-T variant alleles, particularly on the double A29/AW19 background (figure 3A). We found the highest risk to be the combination of rs10044354-TT and two copies of Aw19 with 12 cases and 34 controls (OR=9.9 [4.4-21.2], p=1.66e-07, supplementary table 3). A similar analysis of the *ERAP1*-rs27432 risk haplotype, our top non-MHC association, stratified by single (A29/-) versus double (A29/AW19) Aw19 background, yielded the same trend of increased risk with additional *ERAP1*-rs27432-G variant alleles, particularly on the double A29/AW19 background (OR=6.2 [2.7-15.51], p=1.54e-06, figure 3B and supplementary table 4). We next calculated the combined effects of the *ERAP1* risk haplotype tagged by rs27432, and the *ERAP2* risk haplotype tagged by rs10044354 (figure 3C). We found that the highest risk is conferred by the combination of *ERAP1*-rs27432-GG and *ERAP2*-rs10044354-TT (OR=3.6 [1.62-9.45], p=4.03e-04, supplementary table 5), and as mentioned above, our data are consistent with additive effects of the variants/haplotypes. We next combined all risk haplotypes to a single risk analysis. Due to the small number of cases, we combined the genotypes of intermediate genotypes into four main groups: 1) homozygous to the protective alleles in both *ERAP1* and *ERAP2*, homozygous in one and 2) heterozygous in the other, 3) homozygous risk allele in either *ERAP1* or *ERAP2*, and 4) homozygous risk allele in both *ERAP1* or *ERAP2* (figure 3D and supplementary table 6). We find a gradual increase in risk with the addition of each risk allele, with the highest risk presented when carrying homozygous risk alleles in both ERAP1 and ERAP2, on top of two copies of A19 alleles (OR=13.53 [3.79-54.77], p=1.17e-05). These results suggest that both *ERAP1* and/or *ERAP2* confer greater BSCR risk, which is further increased in the double Aw19 background.

**Figure 3.**
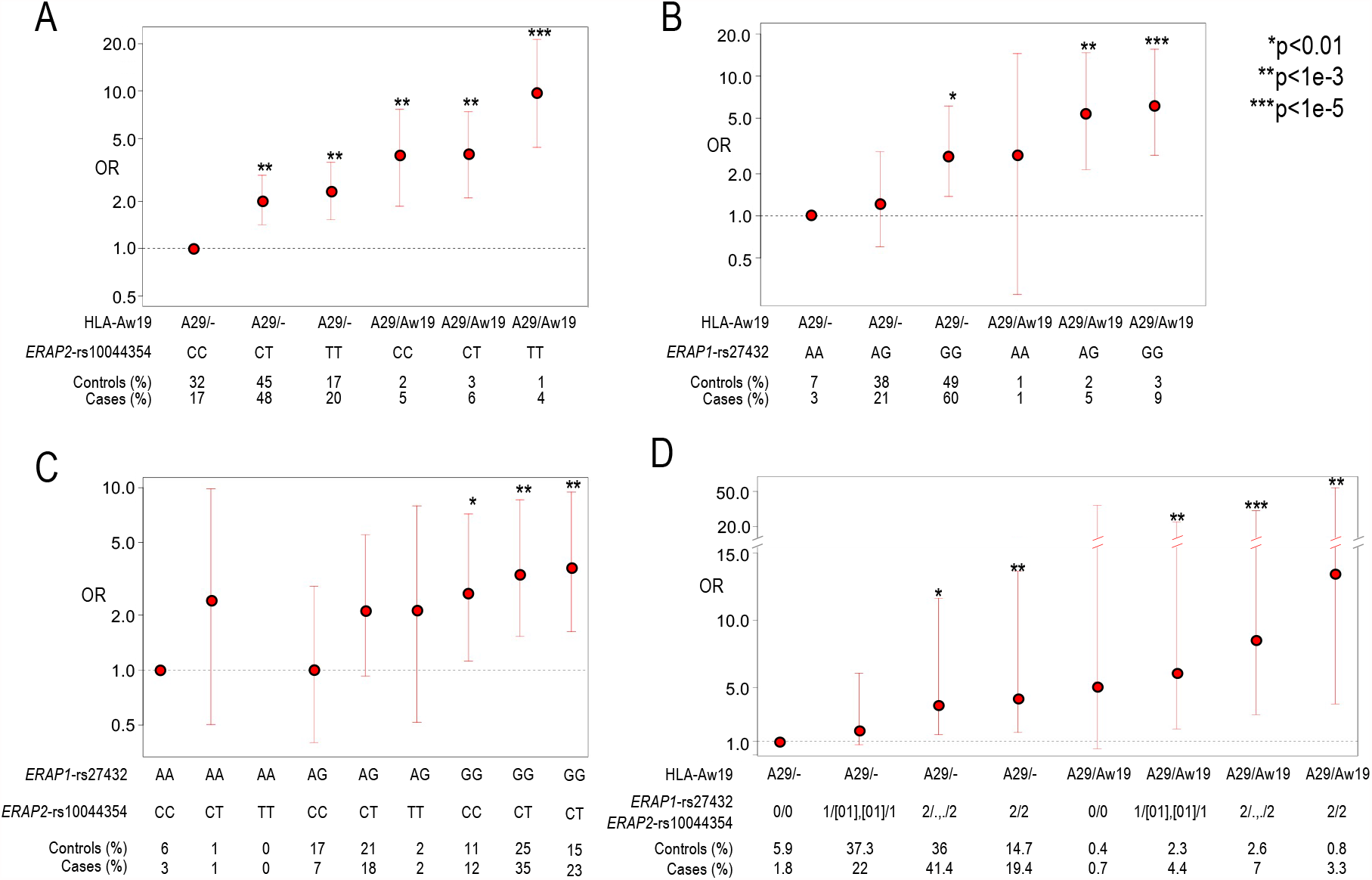
The combined risk of *ERAP1, ERAP2* and two copies of Aw19. Utilizing 286 Birdshot cases and 4,014 controls from GHS cohort #1 to calculate additive risk while combining risk factors in *ERAP1, ERAP2* and Aw19. **A)** An additive genotype model of *ERAP2* risk signal tagged by rs10044354 and single (A29/-) or double (A29/Aw19) Aw19 copies relative to lowest risk combination of rs10044354-CC and one copy of Aw19 allele (A29). **B)** An additive genotype model of *ERAP1* risk signal tagged by rs27432 and single (A29/-) or double (A29/Aw19) Aw19 copies relative to lowest risk combination of rs27432 -AA and one copy of Aw19 allele (A29). **C)** An additive genotype model of *ERAP1* risk signal tagged by rs27432 and *ERAP2* signal tagged by rs10044354 relative to lowest risk combination of rs27432-AA and rs10044354-CC. **D)** An additive genotype model of *ERAP1* and *ERAP2* risk signals and single (A29/-) or double (A29/Aw19) Aw19 copies relative to lowest risk combination. The genotypes are combined as following: 0 = *ERAP1* and *ERAP2* homozygous for protective allele. 1/[01],[01]/1 = either homozygous protective or heterozygous genotypes of both *ERAP1* and *ERAP2*. 2/.,./2 = homozygous risk allele of either *ERAP1* or *ERAP2*. 2/2 = homozygous risk allele of both *ERAP1* and *ERAP2*.

### Absolute BSCR risk

We calculated what would be the absolute risk of BSCR when considering all risk alleles as presented in figure 3D. Since the prevalence of BSCR in general population is estimated at 0.2- 1.7:100,000 ^1^, we used 1:100,000 as an approximation. We further calculated the absolute risk when carrying one A29 carrier based on the frequency of A29 in UKB EUR population of 8%, and reach an absolute risk of 1:29,000 (supplementary table 6). We find that the absolute risk climbs with each risk genotype presented in figure 3D, reaching the most prominent risk at 1:2,160 for cases that carry homozygous risk alleles for both ERAP1 and ERAP2, and two copies of Aw19 alleles. Exhibiting a significant increase in absolute risk of disease when carrying all three risk haplotypes.

## Discussion

The sequencing of a new large BSCR patient cohort and HLA-A29 controls has confirmed the importance of the *ERAP1* and *ERAP2* polymorphisms in increasing risk for developing BSCR. *ERAP1* and *ERAP2* reside back-to-back on chromosome five in opposite orientation and share the regulatory regions, which upregulate one and downregulate the other, and vice versa^15^. The association of both *ERAP1* and *ERAP2* haplotypes is consistent with a mechanism in which coordinated decreased *ERAP1* and increased *ERAP2* expression contribute to disease risk. Several studies have reported that the *ERAP1* and *ERAP2* haplotypes affect their expression as well as the resulting peptidome^13,15,32^

We report the novel finding that several other HLA-Aw19 family alleles (HLA-A29, A30, A31, A33) contribute additional risk as the second HLA-A allele, in addition to the established HLA-A29 risk allele. HLA-Aw19 family alleles have a similar antigen-binding sequence and therefore would bind similar peptide motifs^32^. Hence, the enrichment of Aw19 alleles in cases supports the inferred mechanism underlying activation of the immune response in BSCR: having two copies of these alleles may increase the cell-surface presentation of specific types of peptides in BSCR cases compared to HLA-A29 positive controls. Furthermore, we found that the HLA-A32 allele within the Aw19 family is potentially protective.

Our results indicate that a decreased expression of ERAP1 and an increased expression of ERAP2 confer stronger risk for BSCR than each one separately. Furthermore, this effect is increased in the presence of two copies of HLA-Aw19. The combined and additive effect of risk factors associated with peptide processing and presentation is suggestive of a peptide presentation threshold hypothesis as a driving mechanism for the immune response underlying development of BSCR disease (Figure 4). Results from this and other studies suggest that increased ERAP2 along with decreased ERAP1 expression in BSCR cases would lead to higher availability of ERAP2-processed peptides for presentation onto HLA class I proteins. Additional HLA-Aw19 alleles, with similar peptide-binding properties, would increase presentation of similar peptides. Therefore, both the production of a unique peptide pool by dominant ERAP2 activity and the increased expression of HLA-Aw19 risk allele proteins presenting these peptides may increase the likelihood that a putative ocular autoantigen would be processed and presented above a certain threshold to activate an immune response (Figure 4, left panel). On the other hand, having lower expression of ERAP2 (and higher expression of ERAP1), along with a single HLA-A29 allele, lowers the ocular antigenic peptide presentation below the threshold and thus reduces the risk of generating the immunological response leading to BSCR in HLA-A29 healthy control carriers (Figure 4, right panel). This further highlights the importance of the shaping and generation of the available peptide pool by ERAPs to be presented by specific HLA class I proteins in promoting the generation of an immune response or, in the case of autoimmune disease, an aberrant response to a self-antigen.

**Figure 4:**
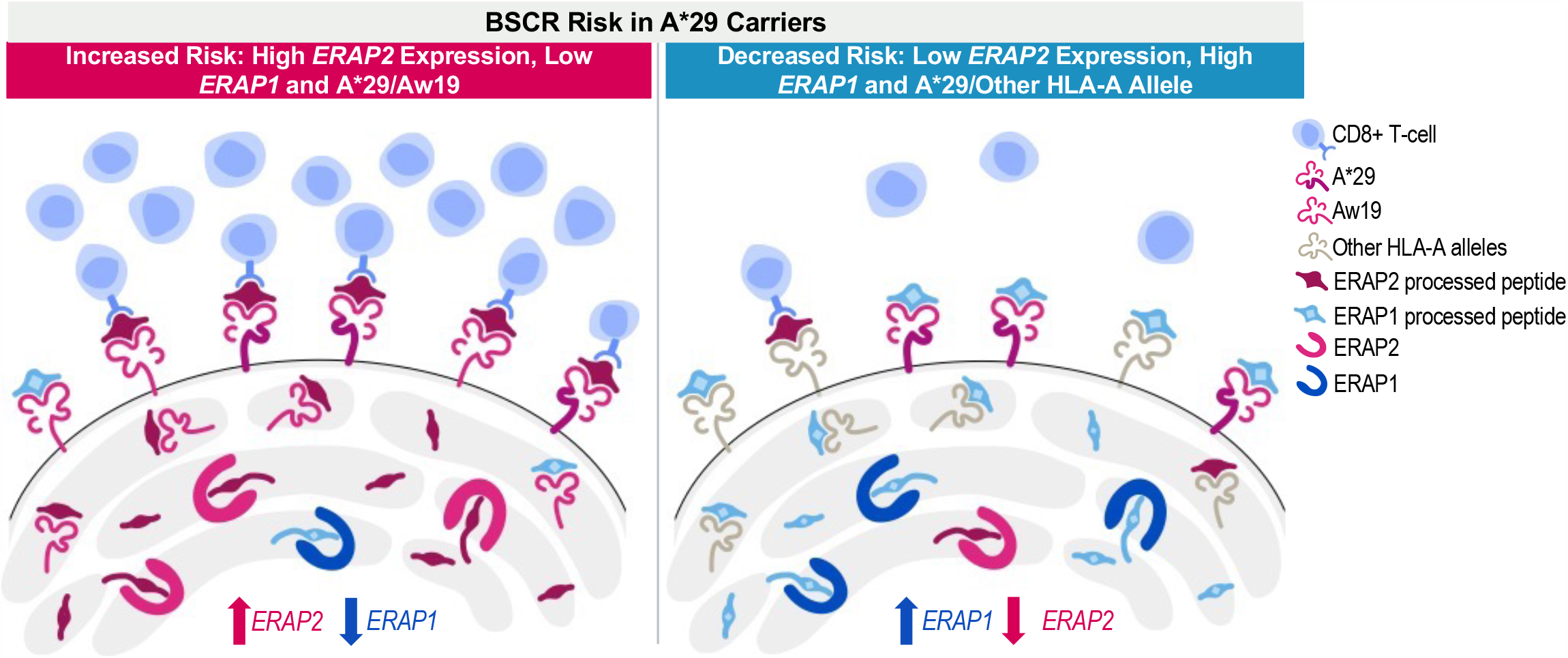
Peptide presentation threshold mechanism in BSCR. An increased expression of *ERAP2* and a decreased expression of *ERAP1*, in addition to two copies of Aw19 alleles, increases the risk for BSCR (left), while a decreased expression of *ERAP2* and an increased expression of *ERAP1*, in presence of one copy of the A29 allele, decreases the risk for BSCR (right).

ERAP1 and ERAP2 polymorphisms and risk haplotypes have also been reported in other HLA class I-associated autoimmune diseases ^35,36^. Polymorphisms in *ERAP1* increase risk for Ankylosing Spondylitis in HLA-B*27 carriers, for psoriasis vulgaris in HLA-C*06 carriers, and for Behçet’s disease in HLA-B*51 carriers, further supporting the combinatorial impact of peptide trimming and HLA class I allele in initiating autoimmune responses ^37-40^. Ankylosing Spondylitis and Behçet’s disease-associated ERAP1 variants have also been experimentally shown to shape the resulting HLA-B*27 and HLA-B*51 peptidome, respectively ^41,42^. Therefore, it is possible that the combination of risk ERAP1/ERAP2 haplotypes and specific risk HLA class I alleles can predispose an individual to develop an HLA class I associated disease in a similar fashion as we hypothesize for BSCR. This implies that the peptide threshold hypothesis may have broader implications as a disease mechanism in HLA class I associated immunological diseases

HLA-A32 is the only HLA-Aw19 member that is found at lower rates in BSCR patients compared to controls, suggesting that it could be protective. The HLA-Aw19 serotype was initially identified by antibody binding to related family members; however, this identifies the HLA-A proteins based on structure outside of the peptide-binding groove. Serofamilies have since been re-analyzed by overall and peptide binding region sequences ^32^. Comparison of the sequences in the peptide binding region reveals that HLA-A32 is more distantly related than the other Aw19 alleles which are identified as novel risk factors in this present study: HLA-A29, A30, A31, A33. When examining the differences in sequence between these Aw19 alleles, two main differences are evident: at position 9, which is part of the peptide binding domain, and a stretch of amino-acids at positions 79-83 that is only found in HLA-A32 and not the other Aw19 alleles (supplementary figure 1). Theoretically, the peptide pool bound by HLA-A32 would differ from the remaining members of the Aw19 family and would not activate the same subset of responding CD8 T cells. This adds further evidence supporting the hypothesis of the threshold requirement of an increased concentration of the driving autoantigenic peptide pool presented on high-risk HLA-A proteins as a driving component for development of BSCR uveitis.

HLA class I alleles not only present peptides to CD8 T cells which can initiate an adaptive immune response, but are also recognized by killer immunoglobulin receptors (KIRs) on NK cells involved in innate immune surveillance. The amino acids at positions 79-83 codes in HLA-A32 contain the Bw4 epitope, shared by a subset of HLA-B and HLA-A alleles known to bind KIR3DL1, which delivers an inhibitory signal to the NK cell^43^. Interestingly, HLA-A32 is the only allele in the Aw19 serotype family that contains an HLA-Bw4 motif and thus is a ligand for the inhibitory KIR3DL1. There are reports of autoimmune HLA class I associated diseases with protective KIR3DL1 or Bw4 genetic associations^44-47^.

In summary, the combinatorial impact of ERAP1/2 shaping the immunopeptidome along with differential peptide selection by the key residues in HLA-A29 and HLA-Aw19 family members supports the immunological hypothesis of a peptide pool that is generated by this combination and available for immune cell recognition and activation initiating an inflammatory cascade. Avenues to reduce the expression and recognition of ERAP2-processed and HLA-Aw19-presented peptides in the eye may be beneficial against BSCR disease and/or progression.

## Data Availability

The data supporting the findings of this manuscript are reported in the main text, in the figures, or in the supplementary materials.

## Acknowledgements

We thank Jonas Kuiper for his expertise and assistance providing results from previous BSCR genome-wide association analysis, as well as performing additional QC and fine-mapping. We also thank Rachel Kirschner for her expertise constructing the scientific illustration pertaining to the hypothesis of quantitative threshold of peptide presentation.

## Abbreviations

Aw19: HLA-Aw’19 broad antigen serotype group
BSCR: Birdshot Chorioretinopathy
GHS: Geisinger Health System
MAF: minor allele frequency
UKB: UK Biobank

## References

1 Minos, E. et al. Birdshot chorioretinopathy: current knowledge and new concepts in pathophysiology, diagnosis, monitoring and treatment. Orphanet J Rare Dis 11, 61, doi:10.1186/s13023-016-0429-8 (2016).

2 Shao, E. H., Menezo, V. & Taylor, S. R. Birdshot chorioretinopathy. Curr Opin Ophthalmol 25, 488– 494, doi:10.1097/ICU.0000000000000101 (2014).

3 Menezo, V. & Taylor, S. R. Birdshot uveitis: current and emerging treatment options. Clin Ophthalmol 8, 73–81, doi:10.2147/OPTH.S54832 (2014).

4 Ryan, S. J. & Maumenee, A. E. Birdshot retinochoroidopathy. Am J Ophthalmol 89, 31–45, doi:10.1016/0002-9394(80)90226-3 (1980).

5 Nussenblatt, R. B., Mittal, K. K., Ryan, S., Green, W. R. & Maumenee, A. E. Birdshot retinochoroidopathy associated with HLA-A29 antigen and immune responsiveness to retinal S-antigen. Am J Ophthalmol 94, 147–158 (1982).

6 Levinson, R. D., Brezin, A., Rothova, A., Accorinti, M. & Holland, G. N. Research criteria for the diagnosis of birdshot chorioretinopathy: results of an international consensus conference. Am J Ophthalmol 141, 185–187, doi:10.1016/j.ajo.2005.08.025 (2006).

7 Brezin, A. P., Monnet, D., Cohen, J. H. & Levinson, R. D. HLA-A29 and birdshot chorioretinopathy. Ocul Immunol Inflamm 19, 397–400, doi:10.3109/09273948.2011.619295 (2011).

8 Kuiper, J. J. et al. A genome-wide association study identifies a functional ERAP2 haplotype associated with birdshot chorioretinopathy. Hum Mol Genet 23, 6081–6087, doi:10.1093/hmg/ddu307 (2014).

9 LeHoang, P. et al. HLA-A29.2 subtype associated with birdshot retinochoroidopathy. Am J Ophthalmol 113, 33–35, doi:10.1016/s0002-9394(14)75749-6 (1992).

10 Herbort, C. P., Jr. et al. Why birdshot retinochoroiditis should rather be called ‘HLA-A29 uveitis’? Br J Ophthalmol 101, 851–855, doi:10.1136/bjophthalmol-2016-309764 (2017).

11 Kuiper, J. J. et al. Detection of choroid-and retina-antigen reactive CD8(+) and CD4(+) T lymphocytes in the vitreous fluid of patients with birdshot chorioretinopathy. Hum Immunol 75, 570–577, doi:10.1016/j.humimm.2014.02.012 (2014).

12 Pulido, J. S. et al. Histological findings of birdshot chorioretinopathy in an eye with ciliochoroidal melanoma. Eye (Lond) 26, 862–865, doi:10.1038/eye.2012.10 (2012).

13 Kuiper, J. J. W. et al. Functionally distinct ERAP1 and ERAP2 are a hallmark of HLA-A29-(Birdshot) Uveitis. Hum Mol Genet 27, 4333–4343, doi:10.1093/hmg/ddy319 (2018).

14 Lopez de Castro, J. A. How ERAP1 and ERAP2 Shape the Peptidomes of Disease-Associated MHC-I Proteins. Front Immunol 9, 2463, doi:10.3389/fimmu.2018.02463 (2018).

15 Paladini, F. et al. An allelic variant in the intergenic region between ERAP1 and ERAP2 correlates with an inverse expression of the two genes. Sci Rep 8, 10398, doi:10.1038/s41598-018-28799-8 (2018).

16 . (!!! INVALID CITATION !!! 14,15).

17 Alvarez-Navarro, C., Martin-Esteban, A., Barnea, E., Admon, A. & Lopez de Castro, J. A. Endoplasmic Reticulum Aminopeptidase 1 (ERAP1) Polymorphism Relevant to Inflammatory Disease Shapes the Peptidome of the Birdshot Chorioretinopathy-Associated HLA-A*29:02 Antigen. Mol Cell Proteomics 14, 1770–1780, doi:10.1074/mcp.M115.048959 (2015).

18 Andres, A. M. et al. Balancing selection maintains a form of ERAP2 that undergoes nonsense-mediated decay and affects antigen presentation. PLoS Genet 6, e1001157, doi:10.1371/journal.pgen.1001157 (2010).

19 Sarkizova, S. et al. A large peptidome dataset improves HLA class I epitope prediction across most of the human population. Nat Biotechnol 38, 199–209, doi:10.1038/s41587-019-0322-9 (2020).

20 The Standardization Of Uveitis Nomenclature Sun Working, G. et al. Classification criteria for birdshot chorioretinitis. Am J Ophthalmol, doi:10.1016/j.ajo.2021.03.059 (2021).

21 Van Hout, C. V. et al. Exome sequencing and characterization of 49,960 individuals in the UK Biobank. Nature 586, 749–756, doi:10.1038/s41586-020-2853-0 (2020).

22 Reid, J. G. et al. Launching genomics into the cloud: deployment of Mercury, a next generation sequence analysis pipeline. BMC Bioinformatics 15, 30, doi:10.1186/1471-2105-15-30 (2014).

23 Bai, Y., Ni, M., Cooper, B., Wei, Y. & Fury, W. Inference of high resolution HLA types using genome-wide RNA or DNA sequencing reads. BMC Genomics 15, 325, doi:10.1186/1471-2164-15-325 (2014).

24 Robinson, J., Soormally, A. R., Hayhurst, J. D. & Marsh, S. G. E. The IPD-IMGT/HLA Database - New developments in reporting HLA variation. Hum Immunol 77, 233–237, doi:10.1016/j.humimm.2016.01.020 (2016).

25 Jia, X. et al. Imputing amino acid polymorphisms in human leukocyte antigens. PLoS One 8, e64683, doi:10.1371/journal.pone.0064683 (2013).

26 Rich, S. S. et al. The Type 1 Diabetes Genetics Consortium. Ann N Y Acad Sci 1079, 1–8, doi:10.1196/annals.1375.001 (2006).

27 Delaneau, O., Zagury, J. F., Robinson, M. R., Marchini, J. L. & Dermitzakis, E. T. Accurate, scalable and integrative haplotype estimation. Nat Commun 10, 5436, doi:10.1038/s41467-019-13225-y (2019).

28 Das, S. et al. Next-generation genotype imputation service and methods. Nat Genet 48, 1284–1287, doi:10.1038/ng.3656 (2016).

29 Mbatchou, J. et al. Computationally efficient whole genome regression for quantitative and binary traits. bioRxiv, 2020.2006.2019.162354, doi:10.1101/2020.06.19.162354 (2020).

30 Zhou, W. et al. Efficiently controlling for case-control imbalance and sample relatedness in large-scale genetic association studies. Nat Genet 50, 1335–1341, doi:10.1038/s41588-018-0184-y (2018).

31 Purcell, S. et al. PLINK: a tool set for whole-genome association and population-based linkage analyses. Am J Hum Genet 81, 559–575, doi:10.1086/519795 (2007).

32 McKenzie, L. M., Pecon-Slattery, J., Carrington, M. & O’Brien, S. J. Taxonomic hierarchy of HLA class I allele sequences. Genes Immun 1, 120–129, doi:10.1038/sj.gene.6363648 (1999).

33 Coulombe-Huntington, J., Lam, K. C., Dias, C. & Majewski, J. Fine-scale variation and genetic determinants of alternative splicing across individuals. PLoS Genet 5, e1000766, doi:10.1371/journal.pgen.1000766 (2009).

34 Sanz-Bravo, A. et al. Allele-specific Alterations in the Peptidome Underlie the Joint Association of HLA-A*29:02 and Endoplasmic Reticulum Aminopeptidase 2 (ERAP2) with Birdshot Chorioretinopathy. Mol Cell Proteomics 17, 1564–1577, doi:10.1074/mcp.RA118.000778 (2018).

35 Babaie, F. et al. The roles of ERAP1 and ERAP2 in autoimmunity and cancer immunity: New insights and perspective. Mol Immunol 121, 7–19, doi:10.1016/j.molimm.2020.02.020 (2020).

36 Yao, Y., Liu, N., Zhou, Z. & Shi, L. Influence of ERAP1 and ERAP2 gene polymorphisms on disease susceptibility in different populations. Hum Immunol 80, 325–334, doi:10.1016/j.humimm.2019.02.011 (2019).

37 Evans, D. M. et al. Interaction between ERAP1 and HLA-B27 in ankylosing spondylitis implicates peptide handling in the mechanism for HLA-B27 in disease susceptibility. Nat Genet 43, 761–767, doi:10.1038/ng.873 (2011).

38 Wisniewski, A. et al. The association of ERAP1 and ERAP2 single nucleotide polymorphisms and their haplotypes with psoriasis vulgaris is dependent on the presence or absence of the HLA-C*06:02 allele and age at disease onset. Hum Immunol 79, 109–116, doi:10.1016/j.humimm.2017.11.010 (2018).

39 Genetic Analysis of Psoriasis, C. et al. A genome-wide association study identifies new psoriasis susceptibility loci and an interaction between HLA-C and ERAP1. Nat Genet 42, 985–990, doi:10.1038/ng.694 (2010).

40 Takeuchi, M. et al. A single endoplasmic reticulum aminopeptidase-1 protein allotype is a strong risk factor for Behcet’s disease in HLA-B*51 carriers. Ann Rheum Dis 75, 2208–2211, doi:10.1136/annrheumdis-2015-209059 (2016).

41 Sanz-Bravo, A. et al. Ranking the Contribution of Ankylosing Spondylitis-associated Endoplasmic Reticulum Aminopeptidase 1 (ERAP1) Polymorphisms to Shaping the HLA-B*27 Peptidome. Mol Cell Proteomics 17, 1308–1323, doi:10.1074/mcp.RA117.000565 (2018).

42 Guasp, P. et al. The Behcet’s disease-associated variant of the aminopeptidase ERAP1 shapes a low-affinity HLA-B*51 peptidome by differential subpeptidome processing. J Biol Chem 292, 9680–9689, doi:10.1074/jbc.M117.789180 (2017).

43 Stern, M., Ruggeri, L., Capanni, M., Mancusi, A. & Velardi, A. Human leukocyte antigens A23, A24, and A32 but not A25 are ligands for KIR3DL1. Blood 112, 708–710, doi:10.1182/blood-2008-02-137521 (2008).

44 Vendelbosch, S. et al. Study on the protective effect of the KIR3DL1 gene in ankylosing spondylitis. Arthritis Rheumatol 67, 2957–2965, doi:10.1002/art.39288 (2015).

45 Berinstein, J. et al. Association of variably expressed KIR3dl1 alleles with psoriatic disease. Clin Rheumatol 36, 2261–2266, doi:10.1007/s10067-017-3784-5 (2017).

46 Petrushkin, H. et al. KIR3DL1/S1 Allotypes Contribute Differentially to the Development of Behcet Disease. J Immunol 203, 1629–1635, doi:10.4049/jimmunol.1801178 (2019).

47 Levinson, R. D. et al. Combination of KIR and HLA gene variants augments the risk of developing birdshot chorioretinopathy in HLA-A*29-positive individuals. Genes Immun 9, 249–258, doi:10.1038/gene.2008.13 (2008).

48 Schrodinger, LLC. The PyMOL Molecular Graphics System, Version 2.0.4 (2015).

