## Supplementary materials for "ERAP1, ERAP2, and two copies of HLA-Aw19 alleles increase the risk for Birdshot Chorioretinopathy in HLA-A29 carriers"

**Supplementary Material**

**Supplementary Figure and Table Legend**

**Supplementary Figure 1. Main differences between risk Aw19 alleles and A32.** A) Sequence differences between risk Aw19 alleles (red) and protective A32 allele (green). A32 exhibits F at position 9 as is the reference A:01:01 allele, while risk alleles are either T or S at that position. The Bw4 epitope sequence is apparent at positions 79-83 or A32 only. B+C)Protein structure calculated using PyMol v2.0.4^1^ exhibit position 9 (pink) in the peptide binding groove and positions 79-83 (bright blue) on the alpha-1 helix.

**Supplementary Table 1. HLA-A 2^nd^ allele frequencies in the French cohort compared to UKB and GHS EUR A29 carriers.** Alleles belonging to the Aw19 broad antigen group that increase risk are A29, A30, A31 and A33 (red) and A32 exhibits protection (green). A logistic regression test with covariates included for sex and six principal components, calculated based on genetic array data for each analytic set. Results are presented for all HLA-A alleles that have three or more case carriers. Alleles are sorted as in Table 1. *Three principal components.

**Supplementary Table 2: Haplotype analysis of *ERAP1*.** Haplotype analysis examining the association of all eight *ERAP1* haplotypes with the case-control status, showing that Hap1 and Hap2 are strongly associated with protection from BSCR.

**Supplementary Table 3: The combined risk of ERAP2 and Aw19.** Utilizing 286 Birdshot cases and 4,014 controls from GHS cohort #1 to calculate additive risk while combining risk factors in *ERAP2* and Aw19. An additive genotype model of *ERAP2* risk signal tagged by rs10044354 and single (A29/-) or double (A29/Aw19) Aw19 copies relative to lowest risk combination of rs10044354-CC and one copy of Aw19 allele (A29).

**Supplementary Table 4: The combined risk of ERAP1 and Aw19.** Utilizing 286 Birdshot cases and 4,014 controls from GHS cohort #1 to calculate additive risk while combining risk factors in *ERAP1* and Aw19. An additive genotype model of *ERAP1* risk signal tagged by rs27432 and single (A29/-) or double (A29/Aw19) Aw19 copies relative to lowest risk combination of rs27432-AA and one copy of Aw19 allele (A29).

**Supplementary Table 5: The combined risk of ERAP1 and ERAP2.** Utilizing 286 Birdshot cases and 4,014 controls from GHS cohort #1 to calculate additive risk while combining risk factors in *ERAP1* and *ERAP2*. An additive genotype model of *ERAP1* risk signal tagged by rs27432 and *ERAP2* signal tagged by rs10044354 relative to lowest risk combination of rs27432-AA and rs10044354-CC.

**Supplementary Table 6: The combined risk of ERAP1, ERAP2 and Aw19.** Utilizing 286 Birdshot cases and 4,014 controls from GHS cohort #1 to calculate additive risk while combining risk factors in *ERAP1*, *ERAP2* and Aw19. An additive genotype model of *ERAP1* and *ERAP2* risk signals and single (A29/-) or double (A29/Aw19) Aw19 copies relative to lowest risk combination. The genotypes are combined as following: 0 = *ERAP1* and *ERAP2* homozygous for protective allele. 1/[01],[01]/1 = either homozygous protective or heterozygous genotypes of both *ERAP1* and *ERAP2*. 2/.,./2 = homozygous risk allele of either *ERAP1* or *ERAP2*. 2/2 = homozygous risk allele of both *ERAP1* and *ERAP2*.

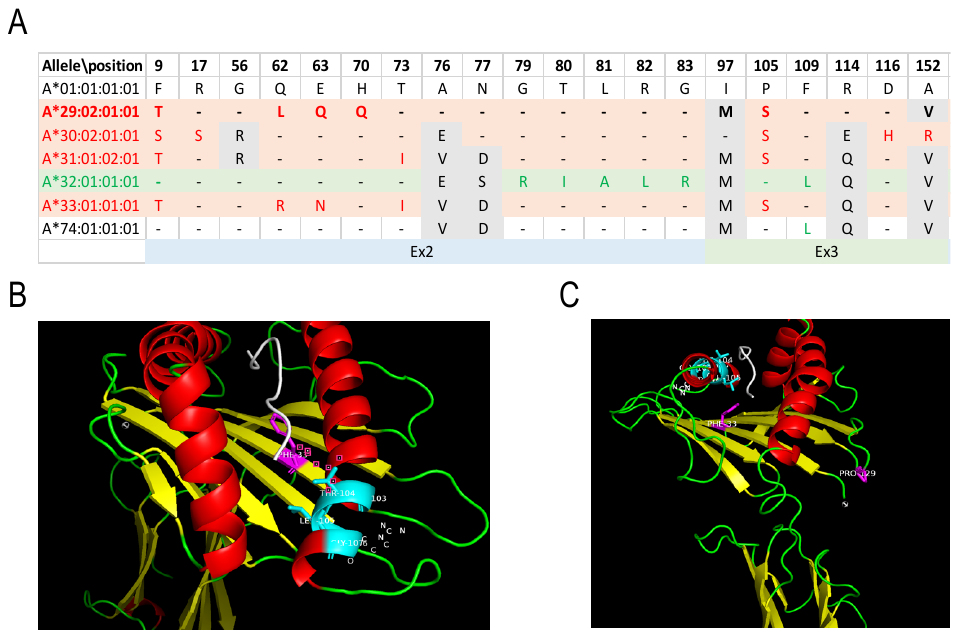

**Supplementary Figure 1. Main differences between risk Aw19 alleles and A32.**

|  | **UParis (A29 EUR Carriers)** | | | | **GHS cohort #1 (A29 EUR)** | | | **GHS cohort #2 (A29 EUR)** | | | **UKB* (A29 EUR)** | | |
| --- | --- | --- | --- | --- | --- | --- | --- | --- | --- | --- | --- | --- | --- |
| **Second HLA-A Allele** | **Cases n=286** | **Ctrls n=108** | **OR (LCI-UCI)** | **p-val** | **Carriers n=4014** | **OR (LCI-UCI)** | **p-val** | **Carriers n=2829** | **OR (LCI-UCI)** | **p-val** | **Carriers n=38,543** | **OR (LCI-UCI)** | **p-val** |
| **A19 co-susceptible (29,30,31,33)** | **41** | **4** | **4.05 (1.40-11.77)** | **0.01** | **247** | **2.21 (1.47-3.33)** | **1.55E-04** | **164** | **2.49 (1.63-3.81)** | **2.34E-05** | **2416** | **2.24 (1.52-3.30)** | **4.57E-05** |
| **A3002** | **13** | **1** | **4.32 (0.55-33.79)** | **0.16** | **45** | **2.78 (1.31-5.88)** | **7.66E-03** | **20** | **4.26 (1.83-9.92)** | **7.68E-04** | **396** | **4.01 (2.05-7.84)** | **4.99E-05** |
| **A3301** | **9** | **0** | **6204540.00 (0.00-Inf)** | **0.98** | **39** | **3.12 (1.29-7.58)** | **0.01** | **32** | **2.69 (1.15-6.28)** | **0.02** | **255** | **2.00 (0.80-5.01)** | **0.14** |
| A0301 | 26 | 15 | 0.71 (0.35-1.45) | 0.35 | 592 | 0.69 (0.45-1.08) | 0.11 | 380 | 0.72 (0.46-1.14) | 0.16 | 5497 | 0.65 (0.43-1.01) | 0.05 |
| A0101 | 71 | 17 | 1.84 (1.01-3.35) | 0.05 | 662 | 1.57 (1.15-2.16) | 0.00 | 448 | 1.59 (1.15-2.19) | 0.01 | 7411 | 1.82 (1.35-2.45) | 0.00 |
| **A3201** | **3** | **4** | **0.22 (0.05-1.04)** | **0.06** | **152** | **0.30 (0.09-0.97)** | **4.38E-02** | **105** | **0.22 (0.06-0.83)** | **2.52E-02** | **1323** | **0.28 (0.08-0.90)** | **3.33E-02** |
| A0201 | 64 | 24 | 0.97 (0.56-1.68) | 0.92 | 1085 | 0.87 (0.64-1.20) | 0.41 | 756 | 0.87 (0.63-1.20) | 0.41 | 10680 | 0.82 (0.61-1.11) | 0.20 |
| **A2902** | **10** | **1** | **3.26 (0.40-26.32)** | **0.27** | **48** | **1.83 (0.80-4.20)** | **0.15** | **37** | **1.94 (0.84-4.46)** | **0.12** | **750** | **2.10 (1.05-4.23)** | **0.04** |
| A2601 | 8 | 3 | 1.09 (0.28-4.26) | 0.90 | 115 | 0.87 (0.39-1.91) | 0.72 | 91 | 0.91 (0.40-2.09) | 0.82 | 834 | 0.73 (0.33-1.61) | 0.44 |
| A2501 | 3 | 2 | 0.55 (0.08-3.61) | 0.54 | 101 | 0.57 (0.16-2.01) | 0.38 | 47 | 0.69 (0.18-2.62) | 0.59 | 673 | 0.66 (0.20-2.15) | 0.49 |
| A2402 | 23 | 11 | 0.85 (0.39-1.85) | 0.67 | 356 | 0.99 (0.61-1.61) | 0.97 | 249 | 1.06 (0.65-1.74) | 0.81 | 2801 | 1.00 (0.63-1.59) | 0.99 |
| **A3101** | **9** | **2** | **1.75 (0.37-8.33)** | **0.48** | **115** | **1.42 (0.68-2.97)** | **0.35** | **75** | **1.57 (0.72-3.41)** | **0.25** | **1015** | **1.24 (0.58-2.64)** | **0.57** |
| A1101 | 14 | 10 | 0.55 (0.23-1.34) | 0.19 | 255 | 0.81 (0.45-1.48) | 0.50 | 177 | 0.83 (0.45-1.53) | 0.54 | 2278 | 0.82 (0.46-1.45) | 0.49 |
| A2301 | 6 | 2 | 1.06 (0.21-5.47) | 0.94 | 94 | 0.85 (0.34-2.17) | 0.74 | 61 | 0.73 (0.28-1.95) | 0.53 | 672 | 0.85 (0.35-2.10) | 0.73 |
| A6801 | 9 | 8 | 0.42 (0.16-1.15) | 0.09 | 128 | 0.95 (0.45-2.04) | 0.90 | 100 | 0.76 (0.35-1.67) | 0.50 | 1142 | 1.43 (0.70-2.91) | 0.33 |
| **A19 all (29,30,31,32,33)** | **44** | **8** | **2.07 (0.92-4.64)** | **0.08** | **399** | **1.49 (1.02-2.18)** | **0.04** | **269** | **1.58 (1.07-2.34)** | **0.02** | **3739** | **1.51 (1.04-2.18)** | **0.03** |

**Supplementary Table 1. HLA-A 2^nd^ allele frequencies in the French cohort compared to UKB and GHS EUR A29 carriers.**

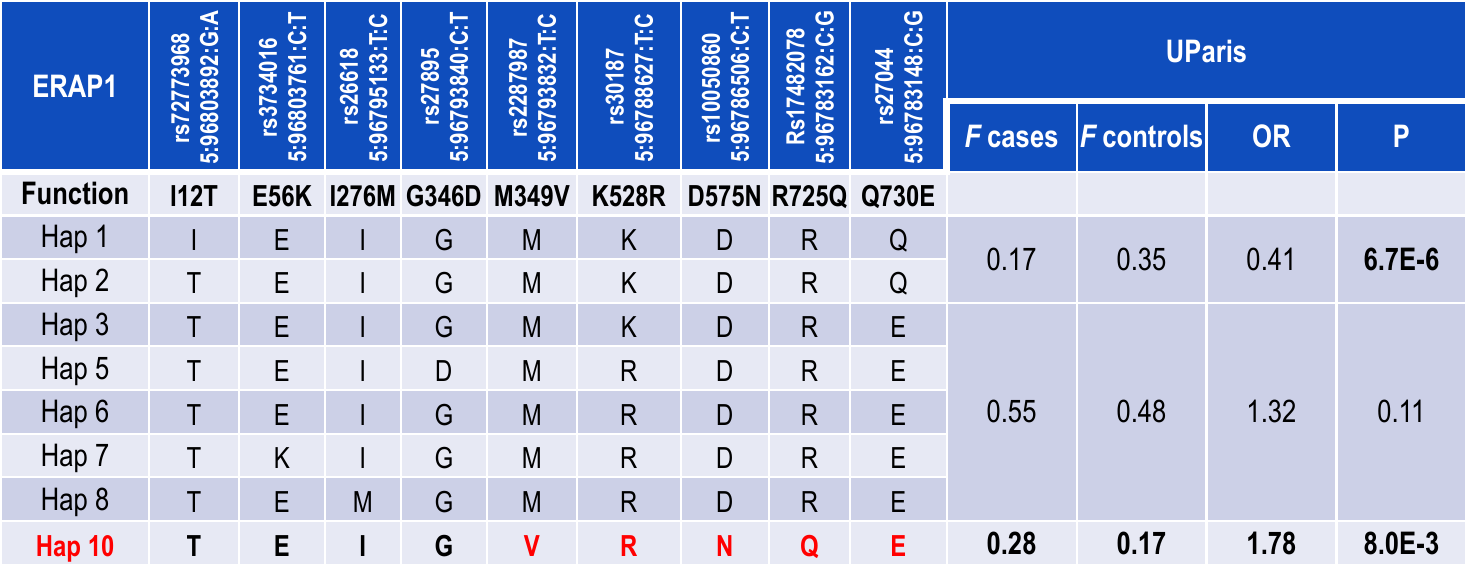

**Supplementary Table 2: Haplotype analysis of *ERAP1*.** Haplotype analysis examining the association of all eight *ERAP1* haplotypes with the case-control status.

| Aw19 | ERAP2- rs10044354 | case_n | contr_n | case_F | controlF | OR | OR  CI_low | OR  CI_high | P |
| --- | --- | --- | --- | --- | --- | --- | --- | --- | --- |
| A29/- | CC | 46 | 1296 | 0.167 | 0.319 | 1.000 | NA | NA | NA |
| A29/- | CT | 132 | 1842 | 0.478 | 0.453 | 2.019 | 1.421 | 2.911 | 3.95E-05 |
| A29/- | TT | 56 | 681 | 0.203 | 0.167 | 2.316 | 1.522 | 3.539 | 4.51E-05 |
| A29/Aw19 | CC | 13 | 94 | 0.047 | 0.023 | 3.891 | 1.861 | 7.644 | 2.13E-04 |
| A29/Aw19 | CT | 17 | 119 | 0.062 | 0.029 | 4.019 | 2.092 | 7.413 | 2.32E-05 |
| A29/Aw19 | TT | 12 | 34 | 0.043 | 0.008 | 9.902 | 4.378 | 21.197 | 1.16E-07 |

**Supplementary Table 3: The combined risk of ERAP2 and Aw19.**

| Aw19 | ERAP1- rs27432 | cases  number | controls  number | cases  freq. | controls  freq. | OR | OR  CI_low | OR  CI_high | P |
| --- | --- | --- | --- | --- | --- | --- | --- | --- | --- |
| A29/- | AA | 9 | 298 | 0.033 | 0.073 | 1.000 | NA | NA | NA |
| A29/- | AG | 58 | 1550 | 0.212 | 0.378 | 1.239 | 0.601 | 2.876 | 0.734 |
| A29/- | GG | 164 | 2001 | 0.601 | 0.488 | 2.713 | 1.374 | 6.105 | 1.72E-03 |
| A29/Aw19 | AA | 2 | 24 | 0.007 | 0.006 | 2.747 | 0.274 | 14.391 | 0.209 |
| A29/Aw19 | AG | 15 | 91 | 0.055 | 0.022 | 5.430 | 2.144 | 14.579 | 9.32E-05 |
| A29/Aw19 | GG | 25 | 133 | 0.092 | 0.032 | 6.196 | 2.708 | 15.515 | 1.54E-06 |

**Supplementary Table 4: The combined risk of ERAP1 and Aw19.**

| ERAP1- rs27432 | ERAP2- rs10044354 | cases  number | controls  number | cases  freq. | controls  freq. | OR | OR_CIlo | OR_CIhi | P |
| --- | --- | --- | --- | --- | --- | --- | --- | --- | --- |
| AA | CC | 7 | 255 | 0.026 | 0.063 | 1.000 | NA | NA | NA |
| AA | CT | 4 | 60 | 0.015 | 0.015 | 2.420 | 0.503 | 9.885 | 0.236 |
| AA | TT | 0 | 2 | 0.000 | 0.000 | 0.000 | 0.000 | 208.116 | 1.000 |
| AG | CC | 19 | 687 | 0.070 | 0.170 | 1.007 | 0.400 | 2.871 | 1.000 |
| AG | CT | 49 | 856 | 0.180 | 0.212 | 2.084 | 0.924 | 5.521 | 0.072 |
| AG | TT | 5 | 86 | 0.018 | 0.021 | 2.113 | 0.515 | 7.964 | 0.197 |
| GG | CC | 32 | 443 | 0.118 | 0.109 | 2.628 | 1.118 | 7.160 | 0.024 |
| GG | CT | 94 | 1030 | 0.346 | 0.255 | 3.323 | 1.525 | 8.594 | 8.29E-04 |
| GG | TT | 62 | 627 | 0.228 | 0.155 | 3.598 | 1.617 | 9.446 | 4.03E-04 |

**Supplementary Table 5: The combined risk of ERAP1 and ERAP2.**

| Aw19 | ERAP1- rs27432 | ERAP2- rs10044354 | cases  number | controls  number | cases  freq. | controls  freq. | OR | OR_CIlo | OR_CIhi | P | Absolute risk |
| --- | --- | --- | --- | --- | --- | --- | --- | --- | --- | --- | --- |
| A29/- | 0 | 0 | 5 | 237 | 0.018 | 0.059 | NA | NA | NA | NA | 3.43E-05 |
| A29/- | 0/1 | 0/1 | 60 | 1510 | 0.220 | 0.373 | 1.883 | 0.752 | 6.071 | 0.197 | 6.45E-05 |
| A29/- | 2_either | 2_either | 113 | 1457 | 0.414 | 0.360 | 3.674 | 1.506 | 11.653 | 1.16E-03 | 1.26E-04 |
| A29/- | 2_both | 2_both | 53 | 596 | 0.194 | 0.147 | 4.211 | 1.666 | 13.665 | 6.26E-04 | 1.44E-04 |
| A29/  Aw19 | 0 | 0 | 2 | 18 | 0.007 | 0.004 | 5.204 | 0.465 | 34.701 | 0.092 | 1.78E-04 |
| A29/  Aw19 | 0/1 | 0/1 | 12 | 93 | 0.044 | 0.023 | 6.079 | 1.929 | 22.651 | 5.23E-04 | 2.08E-04 |
| A29/  Aw19 | 2_either | 2_either | 19 | 104 | 0.070 | 0.026 | 8.604 | 3.002 | 30.314 | 3.09E-06 | 2.95E-04 |
| A29/  Aw19 | 2_both | 2_both | 9 | 31 | 0.033 | 0.008 | 13.527 | 3.791 | 54.766 | 1.17E-05 | 4.63E-04 |

**Supplementary Table 6: The combined risk of ERAP1, ERAP2 and Aw19.**

1	Schrodinger, LLC. *The PyMOL Molecular Graphics System, Version 2.0.4* (2015).

**Regeneron Genetics Center Banner Author List and Contribution Statements**

All authors/contributors are listed in alphabetical order.

**RGC Management and Leadership Team**

Goncalo Abecasis, Aris Baras, Michael Cantor, Giovanni Coppola, Andrew Deubler, Aris Economides, Luca A. Lotta, John D. Overton, Jeffrey G. Reid, Alan Shuldiner, Katia Karalis and Katherine Siminovitch

Contribution: All authors contributed to securing funding, study design and oversight. All authors reviewed the final version of the manuscript.

**Sequencing and Lab Operations**

Christina Beechert, Caitlin Forsythe, M.S., Erin D. Fuller, Zhenhua Gu, M.S., Michael Lattari, Alexander Lopez, M.S., John D. Overton, , Thomas D. Schleicher, M.S., Maria Sotiropoulos Padilla, M.S., Louis Widom, Sarah E. Wolf, M.S., Manasi Pradhan, M.S., Kia Manoochehri, Ricardo H. Ulloa.

Contribution: C.B., C.F., A.L., and J.D.O. performed and are responsible for sample genotyping. C.B, C.F., E.D.F., M.L., M.S.P., L.W., S.E.W., A.L., and J.D.O. performed and are responsible for exome sequencing. T.D.S., Z.G., A.L., and J.D.O. conceived and are responsible for laboratory automation. M.S.P., K.M., R.U., and J.D.O are responsible for sample tracking and the library information management system.

**Genome Informatics**

Xiaodong Bai, , Suganthi Balasubramanian, Boris Boutkov, Gisu Eom, Lukas Habegger, Alicia Hawes, Shareef Khalid, Olga Krasheninina, Rouel Lanche, Adam J. Mansfield, Evan K. Maxwell, Mona Nafde, Sean O’Keeffe, Max Orelus, Razvan Panea, Tommy Polanco, Ayesha Rasool, Jeffrey G. Reid, William Salerno, Jeffrey C. Staples,

Contribution: X.B., A.H., O.K., A.M., S.O., R.P., T.P., A.R., W.S. and J.G.R. performed and are responsible for the compute logistics, analysis and infrastructure needed to produce exome and genotype data. G.E., M.O., M.N. and J.G.R. provided compute infrastructure development and operational support. S.B., S.K., and J.G.R. provide variant and gene annotations and their functional interpretation of variants. E.M., J.S., R.L., B.B., A.B., L.H., J.G.R. conceived and are responsible for creating, developing, and deploying analysis platforms and computational methods for analyzing genomic data.

**Clinical Informatics:**

Michael Cantor, Dadong Li, Deepika Sharma

Contribution: All authors contributed to the clinical informatics of the project

**Research Program Management**

Marcus B. Jones, Jason Mighty, Michelle G. LeBlanc and Lyndon J. Mitnaul

Contribution: All authors contributed to the management and coordination of all research activities, planning and execution. All authors contributed to the review process for the final version of the manuscript.
